# Finding the real COVID-19 case-fatality rates for SAARC countries

**DOI:** 10.1101/2020.10.24.20218909

**Authors:** Md. Rafil Tazir Shah, Tanvir Ahammed, Aniqua Anjum, Anisa Ahmed Chowdhury, Afroza Jannat Suchana

## Abstract

Crude case fatality rate (CFR) is the most accurate when the pandemic is over. Adjustments to the crude CFR measure can better explain the pandemic situation by improving the CFR estimation. However, no study has thoroughly investigated COVID-19 adjusted CFR of the South Asian Association for Regional Cooperation (SAARC) countries. In this study, we estimated both survival interval and underreporting adjusted CFR of COVID-19 for the SAARC countries and observed the CFR changes due to the imposition of fees on COVID-19 tests in Bangladesh. Using the daily records up to 9th October, we implemented a statistical method to remove both the bias in crude CFR, i.e., the delay between disease onset and outcome bias and due to asymptomatic or mild symptomatic cases, reporting rates lower than 50% (95% CI: 10%-50%) bias. According to our findings, Afghanistan had the highest CFR, followed by Pakistan, India, Bangladesh, Nepal, Maldives, and Sri Lanka. Our estimated crude CFR varied from 3.71% to 0.29%, survival interval adjusted CFR varied from 3.77% to 0.3% and further underreporting adjusted CFR varied from 1.1% to 0.08%. We have also found that crude CFR increased from 1.261% to 1.572% after imposing the COVID-19 test fees in Bangladesh. Therefore, the authorities of countries with higher CFR should be looking for strategic counsel from the countries with lower CFR to equip themselves with the necessary knowledge to combat the pandemic. Moreover, caution is needed to report the CFR.

## 1. Introduction

COVID-19 is a highly contagious disease, and the outbreak went global within three months of being first discovered. The disease kept spreading so uncontrollably that even the most adequate healthcare systems around the world were overwhelmed by it. Developing countries are struggling even more [1]. The nature of the disease forced the world to ask questions about the Case Fatality Rate (CFR) of this disease [2]. CFR is an important readout to understand the pandemic severity, and, in the media, CFR is often used to describe the situation regarding COVID-19, as well as any other pandemic. However, during a pandemic, CFR can be misleading [3]. The CFR of a disease is the total number of deaths divided by the total number of cases, i.e., the ratio of fatal cases of a specified condition within a specified time [4]. In CFR calculation during a pandemic, cases might be defined as the total number of confirmed cases, which does not account for the delay between onset of the disease symptoms and outcome, i.e., recovery or death. Therefore, the CFR calculation becomes an underestimate of the actual CFR. By contrast, if we only consider the closed cases where patients have either recovered or died, the real-time CFR estimate remains consistently high throughout [5]. While the crude CFR can give us an approximate idea about the risk of death during the pandemic, it is the most accurate after the pandemic is over [6]. An adjustment to the crude CFR measure can significantly improve the CFR estimates and give us a better idea about the pandemic situation [7]. The Chinese Center for Disease Control and Prevention (China CDC) (2.3%) [8], Lim et al. (approximately 1-2%) [9] and He et al. (2.72% with 95% CI:1.29%□4.16%) [10] estimated the CFR of COVID-19. However, different geographical areas, such as East Asian and Central European countries, differ in the CFR of COVID-19 [11,12]. A study from April 2020 found that the CFR of COVID-19 in Italy was 10.8%, while in Germany, it was just 0.7% [13]. The variation in preventive measures and government policies can be responsible for this difference in CFR [9,14]. For example, about three months into the COVID-19 outbreak, the Bangladesh government inflicted fees on COVID-19 tests on all government labs and hospitals from June 30. Prior to that, all government-run facilities offered COVID-19 tests for free, and 90% of all the whole country’s tests were being conducted on government-controlled sites [15]. The imposition of fees on COVID-19 testing made Bangladesh the only country to do so among all South Asian countries. The Bangladesh government’s official stance was that fees were inflicted to ensure better management and discourage unnecessary tests. Health Experts in Bangladesh believed the imposition of any fee on Covid-19 tests might increase the outbreak size [16–18].

The CFR difference for different countries can provide much-needed information to combatting the pandemic, such as what factors are responsible for speeding up or slowing the outbreak’s progression. Moreover, it will give us a better idea about the fatality rate of COVID-19 of the countries of interest. Therefore, it is of the utmost importance to calculate the CFR of a country with a high degree of representativeness, highlighting the importance of calculating adjusted CFR. However, no study has thoroughly investigated COVID-19 adjusted CFR of SAARC countries, a regional union of eight nations—Afghanistan, Bangladesh, Bhutan, India, Maldives, Nepal, Pakistan, and Sri Lanka. Therefore, this study’s objective was to calculate and compare the COVID-19 CFR for SAARC countries adjusted by the disease’s survival interval and reporting rates. Moreover, we explore the COVID-19 CFR of Bangladesh before and after the test fees imposition.

## 2. Material and methods

We collected the daily record of confirmed cases and deaths attributed to COVID-19 of all member countries of SAARC up to October 9, 2020. Bhutan was not considered in this study since, curiously, no death has been recorded there at all due to COVID-19. Then crude CFR, based on confirmed cases at the time point [*t*], was calculated as

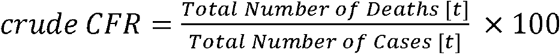

Then we adjusted the crude CFR value by considering the survival interval of COVID-19. As during any point of an ongoing epidemic, the denominator of the crude CFR contains the total number of patients, some of whom may yet fall to their demise due to the disease. The deaths yet to happen cannot possibly be considered in the calculation of CFR. Therefore, we applied a statistical technique to reduce the bias in the crude CFR calculation. The technique considered the uncertainty related to the variability of the interval between disease emergence and death by enabling the probabilistic distribution of the survival interval to vary within a wide range. A gamma distribution with a mean of 13.59 days and a standard deviation of 7.85 days (shape parameter: a=2.99, rate parameter: b=0.22) was used to estimate the adjusted CFR [19]. To allow the mean survival interval to vary between 2 to 6 weeks [20,21], the mean of the Gamma distribution was sampled from a normal distribution with a mean of 13.59 days and a standard deviation of 2 days. Likewise, the standard deviation was sampled from a normal distribution with a mean and a standard deviation of 7.85 days and 1 day, respectively. A maximum likelihood method was then used to estimate the growth rate (r) of the COVID-19 outbreak in the selected SAARC countries. In Monte-Carlo simulations (with 1000 independent replications), Gamma distribution’s moment generating function was used to calculate the correction factor

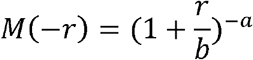

for the adjusted CFR on each calendar day [22]. Finally, we calculated the adjusted CFR using the following formula:

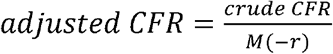

Furthermore, assuming 50% lower reporting rates (95% CI 10%– 50%) of COVID-19 [3,23] due to the asymptomatic cases or exhibition of mild symptoms, we again adjusted the calculated adjusted CFR. The probability of underreporting *(u)* was sampled from a Beta distribution with the shape parameter a = 10 and scale parameter b = 4 as,

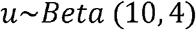

for each Monte-Carlo replication. The parameters of the distribution were selected as such so that the daily reporting rates may vary from 10% to 50%. The 95% confidence interval for *u* was 0.5 - 0.9, while *u* was drawn in the range 0 - 1. The true incidence (*t*) was then estimated by:

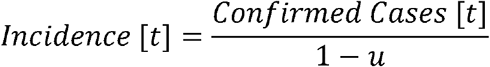

Using the sampled incidence modified for a given probability of low reporting rates, we then again estimated the adjusted CFR.

## 3. Results

Table 1 shows the total confirmed cases and deaths and the confirmation date of the first case of the COVID-19 in the selected SAARC countries. As of 9 October 2020, a total of 7,751,878 confirmed cases of COVID-19, and 120,611 deaths were recorded. India had the highest number of confirmed cases (6,906,151) and deaths (106,490) whereas, Sri Lanka had the lowest numbers (4,488 confirmed cases and 13 deaths) among these countries.

**Table 1.** Overview of the COVID-19 (up to 9 October 2020) situation in the selected SAARC countries.

| Country | First Confirmed Case | Confirmed Cases | Deaths | Population |
| --- | --- | --- | --- | --- |
| Sri Lanka | 28 January 2020 | 4,488 | 13 | 21,413,250 |
| Maldives | 08 March 2020 | 10,742 | 34 | 540,542 |
| Afghanistan | 25 February 2020 | 39,693 | 1,472 | 38,928,341 |
| Nepal | 25 January 2020 | 98,617 | 590 | 29,136,808 |
| Pakistan | 27 February 2020 | 317,595 | 6,552 | 220,892,331 |
| Bangladesh | 09 March 2020 | 374,592 | 5,460 | 164,689,383 |
| India | 30 January 2020 | 6,906,151 | 106,490 | 1,380,004,385 |
| Total |  | 7,751,878 | 120,611 | 1,855,605,040 |

Figure 1 shows the crude CFR, adjusted CFR (accounted for the survival interval), and further adjusted CFR (also considering reporting rates of less than 50%) of the selected South Asian countries on 9 October 2020. In all three scenarios, Afghanistan had the highest CFR, and Sri Lanka had the lowest. The crude CFR varied from 3.71% (Afghanistan) to 0.29% (Sri Lanka), while adjusted CFR varied from 3.77% (95% CI: 3.71%-3.84%) to 0.3% (95% CI: 0.29%-0.30%). When we further adjusted the CFR considering the underreported cases, the CFR varied from 1.1% (95% CI: 1.07%-1.12%) to 0.08% (95% CI: 0.08%-0.09%).

**Figure 1.**
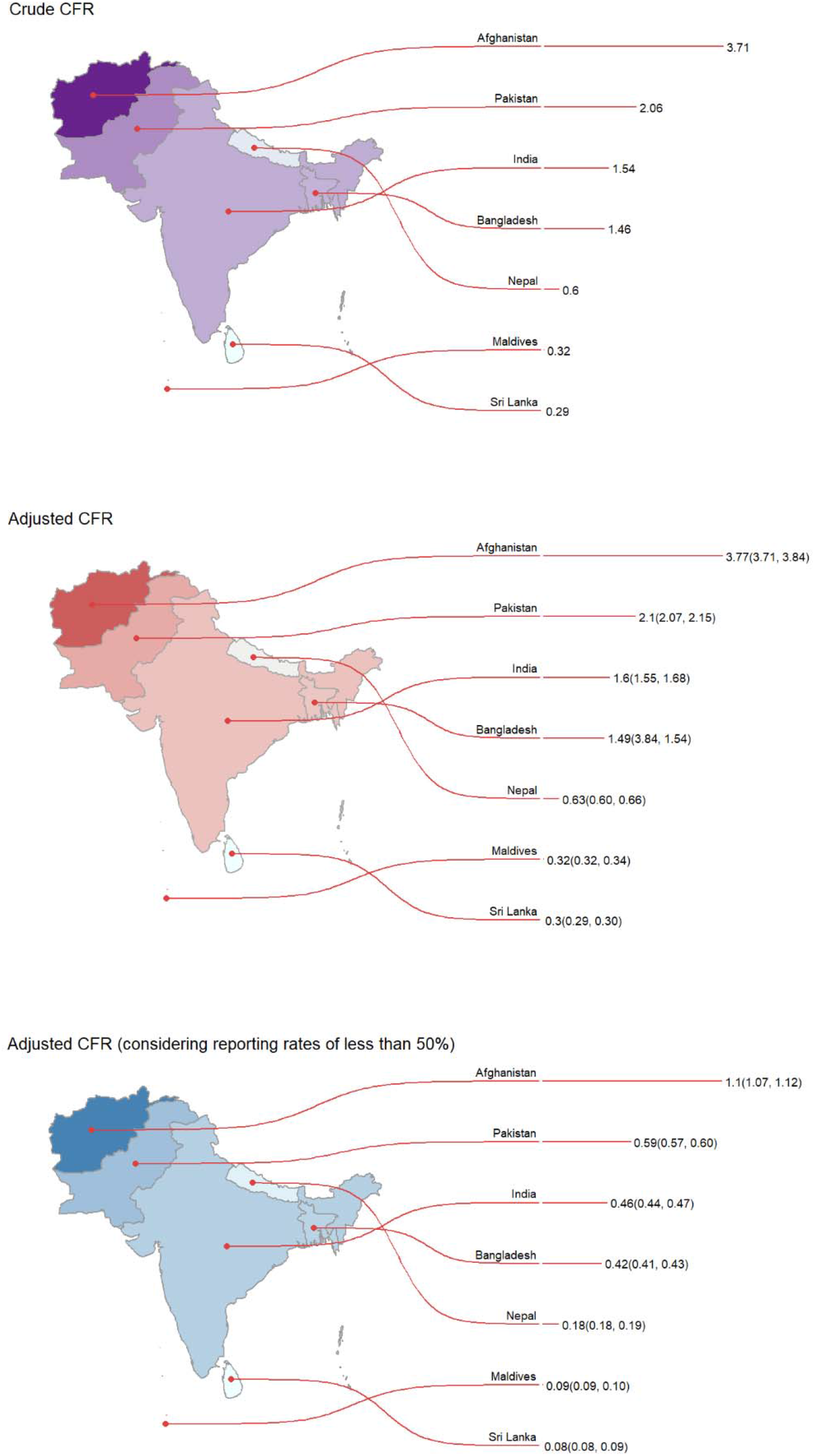
Case fatality rates (95% confidence interval) of the selected SAARC countries (up to 9 October 2020)

Figure 2 and figure 3 indicate that the CFR’s of Maldives, Nepal, and Sri Lanka were relatively low throughout. For Maldives and Nepal, the CFRs were consistent all through the pandemic period, whereas, for Sri Lanka, CFR considerably decreased over time. In Bangladesh, there was a sharp spike in CFR, which became stable over time. Moreover, we found that crude CFR increased from 1.261% to 1.572% after the imposition of the COVID-19 test fees in Bangladesh (Figure 4).

**Figure 2.**
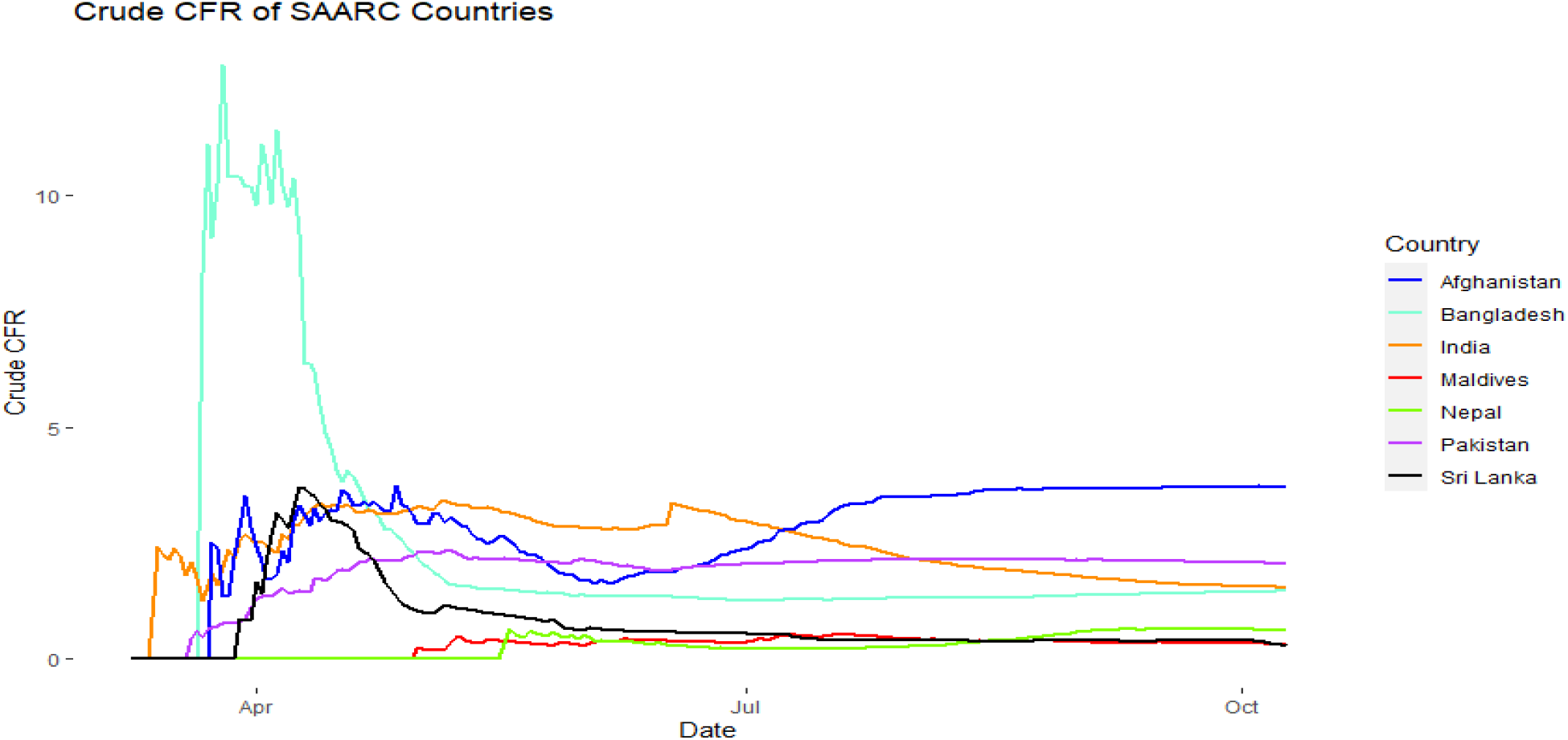
The changes in crude case fatality rates over time for the selected countries.

**Figure 3.**
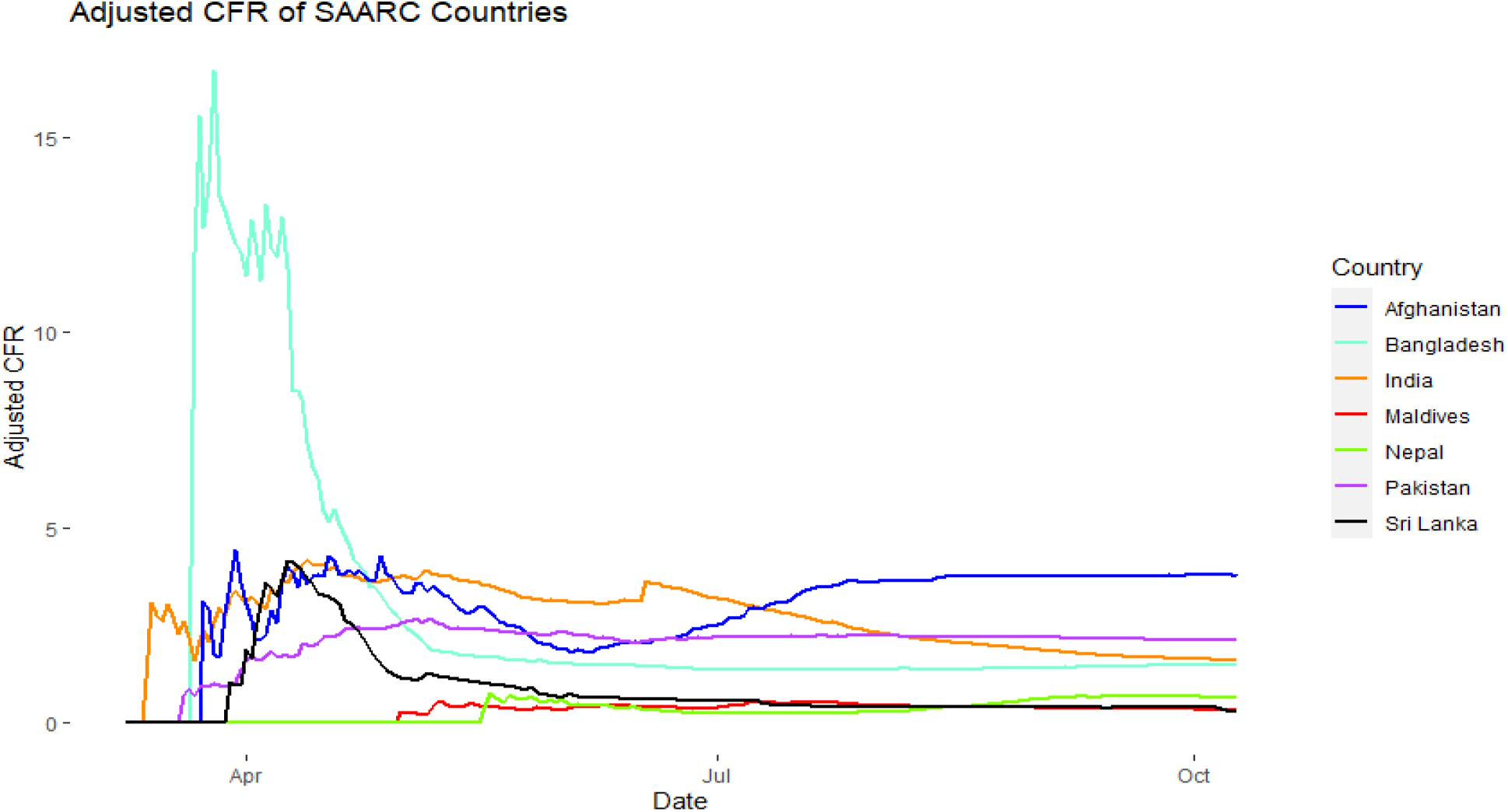
The changes in adjusted case fatality rates over time for the selected countries.

**Figure 4.**
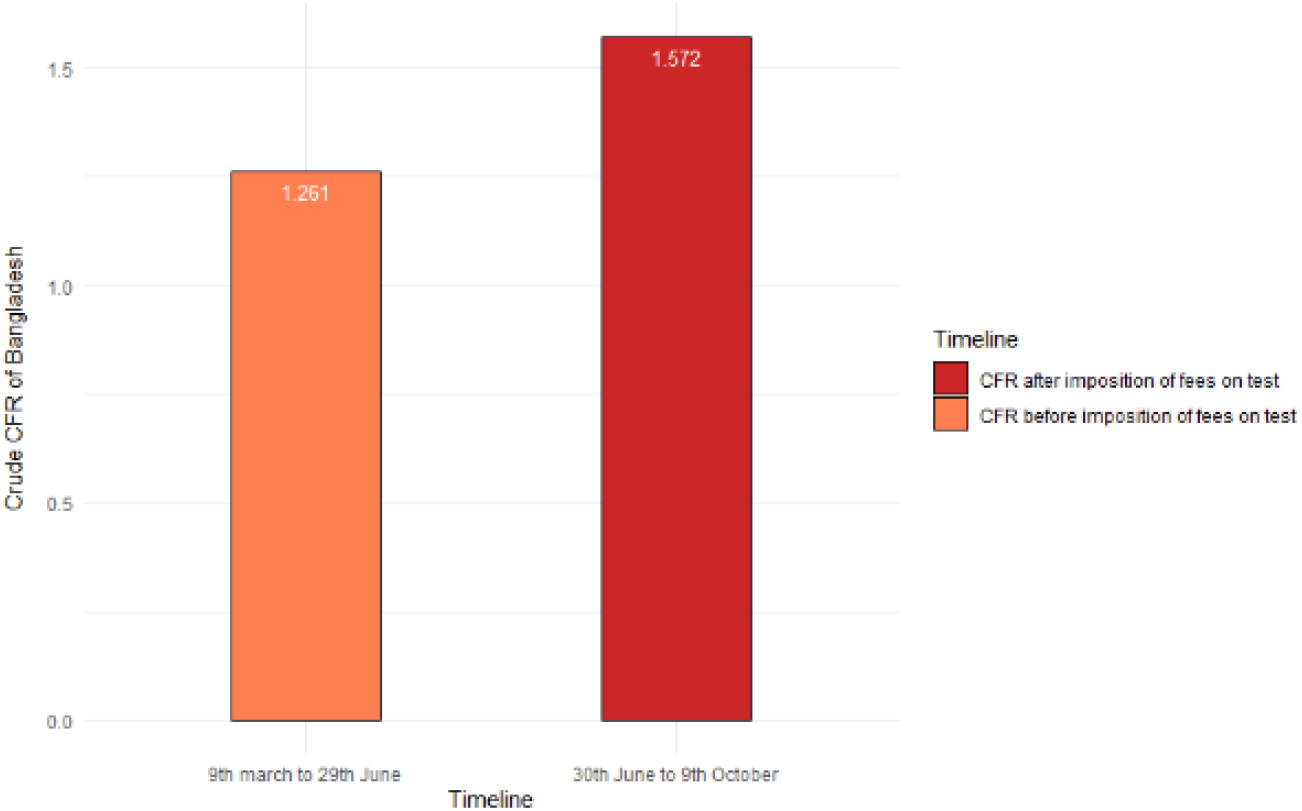
The crude CFR of Bangladesh before and after imposition of fees on COVID-19 tests

## 4. Discussion

Understanding the case fatality rate of the COVID-19 allows policymakers to mitigate the outbreak impact by implementing efficient and effective interventions for disease control. Therefore, in this study, we estimated the adjusted CFR of COVID-19 outbreak for the selected SAARC countries, i.e., Afghanistan, Bangladesh, India, Maldives, Nepal, Pakistan, and Sri Lanka. To our knowledge, this is by far the most comprehensive estimation for the COVID-19 CFR for these selected countries.

There was a difference in estimated CFRs of SAARC countries. Ranking the countries, we observed the highest CFR in Afghanistan, followed by Pakistan, India, Bangladesh, Nepal, Maldives, and Sri Lanka. The variation can be attributed to the public health system, preparedness, and effective interventions of each country. For example, Sri Lanka has a free public health system and has been ranked as 10^th^ on Global Response to Infectious Disease while Bangladesh is 80^th^ [24].

According to our findings, the survival interval adjusted CFR values were slightly greater than that of the respective countries’ crude CFR. The reason is that during an epidemic crude CFR estimation becomes an underestimate of the actual CFR [5]. However, because of both limited numbers of test and presence of asymptotic or mild cases, there exists unreported cases [25–28]. Therefore, after further adjustment for reporting rates lower than 50%, estimated CFRs became less than one-third compared to crude CFR, and survival interval adjusted CFR. In agreement with previous studies [29,30], our estimated CFRs for selected countries were lower than most of the European countries’ CFR. Several factors, such as temperature, the proportion of young people, genetic factors, can be responsible for this variation in CFR [12,14,31–33]. Moreover, we found that CFR of the COVID-19 pandemic is less than SARS, MERS, Bird flu, and Ebola [34,35]. However, as it is highly infectious, and there are many mild or asymptomatic cases, public health concerns must be addressed.

In our estimation, the crude CFR of Bangladesh increased after the imposition of the COVID-19 test fees to discourage unnecessary tests, therefore, ensure better management. As a result, in the first 10 days since the imposition of test fees, there had been a decrease of 8,736 tests in total from the previous 10-day period. More tests can detect more asymptomatic or mild cases, which reduced the mortality rate [12,14]. However, as the decision had affected the poor citizen’s ability or willingness of testing [18], the Bangladesh government decided to cut the test fees by almost half on 20 August [36]. Since the government-imposed fee for COVID-19 is increasing the country’s CFR, immediate steps should be taken to remove the fees so that the tests are affordable to everybody.

## 5. Limitations

The first limitation of this study is that the calculated case-fatality rates refer to the countries’ entire population. Patients with critical health condition, populations with a higher proportion of older adults, inadequate resources and unorganized health care systems can have a higher CFR. [12,37–39]. Second, we could not use country wise mean survival time of COVID-19 patients for the adjusted CFR estimation. Third, we assumed that there were no age-specific and country-specific differences in under-reporting. The children and youths with mild symptoms are tested less often. Moreover, factors such as testing capacity, awareness about the importance of reporting symptoms etc. directly affects the reporting rates of the disease [40]. It is recommended that future researches on similar issues should consider improving on these limitations.

## 6. Conclusion

Survival intervals of the patients and a large number of underreported cases affect the CFR estimation, therefore affecting the countries’ policies. In this regard, the bias-adjusted CFR measure can provide better information to health professionals and policymakers. Therefore, survival interval and underreported cases should be considered while calculating COVID-19 CFR. This will equip us with much better knowledge of the COVID-19 scenario worldwide.

## Data Availability

The data that support the findings of this study are available from the corresponding author, upon reasonable request.

## Acknowledgements

We acknowledge Musaddiqur Rahman Ovi for his support in visualization.

## Conflict of interest statement

There are no conflicts of interest to declare.

## Notes

### Competing Interest Statement

The authors have declared no competing interest.

### Funding Statement

None.

